# Knowledge and Beliefs of General Public of India on COVID-19: A Web-based Cross-sectional Survey

**DOI:** 10.1101/2020.04.22.20075267

**Authors:** Puvvada Rahul Krishna, Krishna Undela, Shilpa Palaksha, Balaji Sathyanarayana Gupta

## Abstract

**Context:** Despite many awareness programs conducted by the governments and other agencies, there are certain false beliefs among the general public of India towards the transmission, prevention, and treatment of COVID-19.

**Aims:** To assess the knowledge and beliefs of the general public of India on COVID-19.

**Materials and Methods:** A web-based cross-sectional survey was conducted between 20th March and 15th April 2020. A 17-item questionnaire was developed, validated, and used for the study. The questionnaire was randomly distributed among the public using Google forms through social media networks. Descriptive analysis was performed to represent the study characteristics, Chi-square test for assessing the associations among the study variables, and logistic regression analysis for identifying the factors influencing the beliefs.

**Results:** A total of 462 participants with a mean (SD) age of 30.66 (11.31) years were responded to the questionnaire. Study participants are having fairly good knowledge of the basic aspects of COVID-19. However, a considerable fraction of participants were having false beliefs towards the transmission of new coronavirus, and prevention & treatment of COVID-19. It was observed that the participants who were aged 31-60 years and >60 years, education level of intermediate or diploma and high school certificate, and occupation as the unskilled workers had more of false beliefs towards COVID-19 compared to their counterparts.

**Conclusion:** Though the overall knowledge on COVID-19 was good enough among the general public of India, still there is a need for education to avoid false beliefs especially among the people who are elderly, having a low level of education, and non-professional workers.

**Key Messages:** A cross-sectional web-based online survey was conducted to assess the knowledge and beliefs of general public of India on COVID-19. It was identified that the knowledge among the general public on COVID-19 is fairly good. However, still there are some false beliefs among the population towards transmission of new coronavirus, and prevention & treatment of COVID-19, especially among the people who are elderly, having low level of education, and non-professional workers.

## Introduction

Coronaviruses are large family of viruses that are known to cause mild to moderate respiratory symptoms. However, the epidemics of two beta coronaviruses namely MERS (Middle East Respiratory Syndrome), SARS (Severe Acute Respiratory Syndrome) has caused over 10,000 deaths over the past two decades.^[1,2]^ In December 2019, scientists had found series of cases with symptoms resemble to pneumonia in Wuhan, China and identified that it was caused by a novel strain of coronavirus called SARS-CoV-2. Later, on 11^th^ February 2020, World Health Organization (WHO) has announced COVID-19 (Coronavirus Disease 2019) as new name of the disease. ^[3]^ The WHO has announced COIVD-19 as pandemic on 11^th^ March 2020. ^[4]^ As of 19^th^ April 2020, 185 countries/regions were affected with SARS-Cov-2 with a total of 2,329,651 confirmed cases and 160,721 deaths. ^[5]^ Of these countries, highest number of cases are seen in United States (735,086 cases), followed by Spain (194,416 cases), Italy (175,925), France (152,978), Germany (143,724) and United Kingdom (115,314). COVID-19 cases in India increasing gradually, and the number of cases as of 19^th^ April 2020 are 16,365 including 521 deaths.^[6]^

Despite general awareness programs conducted by the state and central governments, there are many false beliefs among the general public of India due to circulation of various misconceptions in social media and internet about the spread of novel coronavirus, and prevention & treatment of COVID-19. False beliefs not only misrepresent the situation but may increase the effects of disasters by compromising the public’s collective effort.^[7]^ An advocacy group of BBC said around 46% of adults have been seeing fake news or misleading information about the COVID-19 pandemic and suggested that there is a need to stop the false posts.^[8]^ In Iran, thinking that alcohol can kill the coronavirus, around 600 people died and 3000 were hospitalised after drinking methanol,^[9]^ which is a type of alcohol used in marine, automotive and electricity industries as a renewable energy resource.^[10]^ In a study conducted by Kok et al., it was identified that majority of participants thought government and/or media were exaggerating the situation. ^[11]^

India, with 1.39 billion population, is the second largest internet user in the world with over 560 million internet users, and has over 400 million WhatsApp users as of July 2019. ^[12,13]^ According to the article published in India Today, the Maharashtra cyber cell department has registered 115 cases over spread of fake news since lockdown during COVID-19. ^[14]^ Hence, it is essential to assess the knowledge and beliefs of general public of India towards COVID-19 for taking preventive measures in spreading misconceptions and increase awareness. To the best of our knowledge, this is the first Indian study conducted to assess the knowledge and beliefs of general public of India on COVID-19.

## Material and Methods

A cross-sectional web-based online survey was conducted between 20^th^ March and 15^th^ April 2020. A 17-item questionnaire was developed based on WHO myth busters.^[15]^ The developed questionnaire was validated for relevance, clarity, simplicity and ambiguity by using four point content validity index. ^[16]^ A pilot study was conducted among 15 individuals to identify further barriers in understanding the questionnaire before disseminating the questionnaire. Cronbach’s alpha was used to measure the internal consistency of the questionnaire.^[17]^

The questionnaire consists of three sections. The first section consists of demographics of participants such as age, gender, occupation, education and monthly income. The socioeconomic status of participants were grouped according to modified kuppuswamy scale.^[18]^ The second section has six knowledge based questions focus on knowledge of general public of India on COVID-19. Initial five questions were provided with multiple options with only one correct answer and the last question is about sources of information for which the respondent can choose multiple options. The scoring was given to first five questions. Each correct answer was given score “1” and incorrect answer was given “0”. The third section has 11 questions pertaining to beliefs of people on transmission of new coronavirus, and prevention & treatment of COVID-19. Each question has three options (Yes/No/Don’t Know). The responses ‘Yes and Don’t Know’ were considered as false belief and ‘No’ was considered as correct belief. The correct answer was given score “3”, incorrect “1” and Don’t Know “2”. For the purpose of identifying the factors influencing the knowledge and beliefs, participants were grouped into low and high knowledge and belief score categories based on the cumulative score (≤2 and ≥3 for knowledge, and 11-21 and 22-33 for beliefs).

Using Google forms, the questionnaire was randomly distributed among the public of India including doctors, pharmacists, nurses, medical and engineering students, software employees, retired persons, etc. through emails and social networking platforms such as WhatsApp, LinkedIn, Instagram and Facebook. Also, we requested people to circulate the survey link among their respective family members, friends and colleagues.

This study received an exemption from the Institutional Human Ethics committee of JSS College of Pharmacy, Mysuru as it was considered as no minimal risk research. However, participants were asked to give consent before taking part in the survey.

Mean with standard deviation was calculated for continuous variables and number with percentage was calculated for categorical variables. Chi-square test was used to find the association between demographic details of people and their beliefs on COVID-19, and the factors influencing beliefs were identified by using odds ratio with 95% CI. The results were considered statistically significant with p□0.05. All statistical analyses were performed using Statistical Package for Social Sciences (IBM Corp. Released 2012. IBM SPSS Statistics for Windows, Version 21.0. Armonk, NY, USA: IBM Corp).

## Results

A total of 462 participants with mean (SD) age of 30.66 (11.31) years were responded to the questionnaire. The age of participants ranged from 18 – 74 years. More than half of the responses were received from males (57.58%). Majority of participants were graduates (58.66%) by education and professionals (44.81%) by occupation. Around 50% of participants were from upper middle economic class (Table 1).

**Table 1:** Demographic details of study participants (N = 462)

| Characteristic | Category | No. of Participants (%) |
| --- | --- | --- |
| Age | 18-30 Years | 312 (67.53) |
|  | 31-60 Years | 136 (29.44) |
|  | >60 Years | 14 (3.03) |
| Gender | Female | 196 (42.42) |
|  | Male | 266 (57.58) |
| Education | Postgraduate and above | 125 (27.06) |
|  | Graduate | 271 (58.66) |
|  | Intermediate or Diploma | 46 (9.96) |
|  | High school certificate | 15 (3.25) |
|  | Middle school certificate | 2 (0.43) |
|  | Illiterate | 1 (0.22) |
|  | Prefer not to say | 2 (0.43) |
| Occupation | Professional | 207 (44.81) |
|  | Student or Unemployed | 123 (26.62) |
|  | Self employed | 57 (12.34) |
|  | Skilled worker | 48 (10.39) |
|  | Unskilled worker | 16 (3.46) |
|  | Prefer not to say | 11 (2.38) |
| Monthly Income<br>(in INR) | ≤3,907 | 125 (27.06) |
|  | 3,908–11,707 | 34 (7.36) |
|  | 11,708–19,515 | 47 (10.17) |
|  | 19,516–29,199 | 48 (10.39) |
|  | 29,200 –39,032 | 42 (9.09) |
|  | 39,033–78,062 | 81 (17.53) |
|  | ≥78,063 | 59 (12.77) |
|  | Prefer not to say | 26 (5.63) |
| Socioeconomic<br>scale | Upper (I) | 81 (17.53) |
|  | Upper Middle (II) | 236 (51.08) |
|  | Lower Middle (III) | 36 (7.79) |
|  | Upper Lower (IV) | 108 (23.37) |
|  | Lower (V) | 1 (0.21) |

Overall, the knowledge of participants on COVID-19 is satisfactory. Around 78% of participants were aware of risk factors of COVID-19. Almost all participants (96.96%) knew that the new coronavirus will affect lungs. All participants were aware of symptoms of COVID-19. Majority of participants were aware of modes of transmission of COVID-19 and precautions to be taken if affected with COVID-19. Social media (WhatsApp, Facebook etc), News channels, Government websites, Friends, Family, Neighbours, Healthcare Professionals (Doctors, Pharmacists, Nurses etc.) are the major information sources of these participants. The details of participants responses on knowledge about COVID-19 are presented in table 2.

**Table 2:** Knowledge of study participants on COVID-19 (N = 462)

| **Sl. No.** | **Question** | **Correct answer** | **Correct response n (%)** | **Incorrect response n (%)** |
| --- | --- | --- | --- | --- |
| 1 | What type of people are at the risk of acquiring COVID-19? | All the above | 360 (77.92) | 102 (22.08) |
| 2 | Which body organ will be affected by the new coronavirus? | Lungs | 448 (96.97) | 14 (3.03) |
| 3 | What are the symptoms of COVID-19? | Dry cough, High fever, Sore throat, Difficulty in breathing | 462 (100) | 0 (0.00) |
| 4 | How does the new coronavirus spread? | All the above | 390 (84.42) | 72 (15.58) |
| 5 | What need to be done if a healthy person come in contact with COVID-19 infected person? | Keep away from the infected person and undergo self-quarantine | 457 (98.92) | 5 (1.08) |
| 6 | What is the source of information for your understanding on COVID-19? | Social media (WhatsApp, Facebook, etc.) |  | 210 (45.45) |
|  |  | News channels |  | 275 (59.52) |
|  |  | Government websites |  | 245 (53.03) |
|  |  | Friends, Family, Neighbours |  | 107 (23.16) |
|  |  | Healthcare Professionals (Doctors, Pharmacists, Nurses, etc.) |  | 272 (58.87) |

Majority of participants agreed to the fact that changes in weather (56.49%), hand dryers (70.34%) and taking hot water bath (59.3%) can’t kill the new coronavirus. However, only 42.85% of participants knew the fact that garlic will not help in treating or preventing COVID-19. Majority of participants were aware that the new coronavirus will not spread through mosquito bites (87.22%) and eating non-vegetarian food (88.74%). However, only less than half of the participants knew that UV disinfection lamps (46.53%) and swallowing or gargling warm water or saltwater (41.55%) can’t kill the new coronavirus. Also, majority of participants (88.09%) were aware that the COVID-19 is not the disease of only elderly (Table 3).

**Table 3:** Beliefs of study participants on COVID-19 (N = 462)

| **Sl. No.** | **Question** | **Yes**<br>**n (%)** | **Don't know**<br>**n (%)** | **No**<br>**n (%)** |
| --- | --- | --- | --- | --- |
| 1 | Cold or hot weather can kill the new coronavirus | 73 (15.80) | 128 (27.71) | 261 (56.49) |
| 2 | Taking hot water baths can prevent from getting COVID-19 | 106 (22.94) | 82 (17.74) | 274 (59.3) |
| 3 | New coronavirus can spread through mosquito bites | 15 (3.24) | 44 (9.52) | 403 (87.22) |
| 4 | Hand dryers can kill the new coronavirus | 50 (10.82) | 87 (18.83) | 325 (70.34) |
| 5 | UV (Ultraviolet) disinfection lamps can kill the new coronavirus | 61 (13.2) | 186 (40.25) | 215 (46.53) |
| 6 | If infected with COVID-19, drinking or spraying alcohol and chlorine all over the body can kill the new coronavirus | 80 (17.31) | 77 (16.66) | 305 (66.01) |
| 7 | Swallowing or gargling warm water or saltwater or with bleach can prevent from getting COVID-19 | 182 (39.39) | 88 (19.04) | 192 (41.55) |
| 8 | Ordering products from China will make a person affected to COVID-19 | 91 (19.69) | 98 (21.21) | 273 (59.09) |
| 9 | Eating garlic helps in preventing or treating COVID-19? | 139 (30.08) | 125 (27.05) | 198 (42.85) |
| 10 | The new coronavirus can affect only older people (Age 60 years and above) | 44 (9.52) | 11 (2.38) | 407 (88.09) |
| 11 | Eating non-vegetarian food can cause COVID-19 | 15 (3.24) | 37 (8.01) | 410 (88.74) |

By conducting the logistic regression analysis, it was identified that the age, educational status and occupation were the factors influencing the beliefs of participants on COVID-19. Participants who are aged 31-60 years [OR 1.90 (1.02-3.53); p=0.041] and >60 years [OR 14.67 (4.73-45.50); p<0.001], education level of intermediate or diploma [OR 3.08 (1.08-8.76); p=0.039] and high school certificate [OR 16.71 (4.83-57.86); p<0.001], and occupation as unskilled worker [OR 4.50 (1.41-14.31); p=0.018] had more of false beliefs towards COVID-19 compared to their counterparts (Table 4).

**Table 4:** Factors effecting the beliefs of study participants.

| **Factor** |  | **No. of**<br>**Participants** | **Low Score (11-21)**<br>**n (%)** | **High Score (22-33)**<br>**n (%)** | **Chi<sup>2</sup>**<br>**P-Value** | **OR (95% CI); P-Value** |
| --- | --- | --- | --- | --- | --- | --- |
| Age | 18-30 Years | 312 | 26 (8.33) | 286 (91.67) | <0.001 | Reference |
|  | 31-60 Years | 136 | 20 (14.71) | 116 (85.29) |  | 1.90 (1.02-3.53); 0.041 |
|  | >60 Years | 14 | 8 (57.14) | 6 (42.86) |  | 14.67 (4.73-45.50); <0.001 |
| Gender | Female | 196 | 22 (11.22) | 174 (88.78) | 0.790 | Reference |
|  | Male | 266 | 32 (12.03) | 234 (87.97) |  | 1.08 (0.61-1.93); 0.790 |
| Education | Postgraduate and above | 125 | 8 (6.40) | 117 (93.60) | <0.001 | Reference |
|  | Graduate | 271 | 29 (10.70) | 242 (89.30) |  | 1.75 (0.78-3.95); 0.172 |
|  | Intermediate or Diploma | 46 | 8 (17.39) | 38 (82.61) |  | 3.08 (1.08-8.76); 0.039 |
|  | High school certificate | 15 | 8 (53.33) | 7 (46.67) |  | 16.71 (4.83-57.86); <0.001 |
|  | Middle school certificate | 2 | 1 (50.00) | 1 (50.00) |  | -- |
|  | Illiterate | 1 | 0 (0.00) | 1 (100.00) |  | -- |
|  | Prefer not to say | 2 | 2 (10.00) | 0 (0.00) |  | -- |
| Occupation | Professional | 207 | 19 (9.18) | 188 (90.82) | 0.121 | Reference |
|  | Student or Unemployed | 123 | 13 (10.57) | 110 (89.43) |  | 1.17 (0.56-2.46); 0.680 |
|  | Self employed | 57 | 8 (14.04) | 49 (85.96) |  | 1.62 (0.67-3.91); 0.284 |
|  | Skilled worker | 48 | 8 (16.67) | 40 (83.33) |  | 2.0 (0.81-4.84); 0.129 |
|  | Unskilled worker | 16 | 5 (31.25) | 11 (68.75) |  | 4.50 (1.41-14.31); 0.018 |
|  | Prefer not to say | 11 | 1 (9.09) | 10 (90.91) |  | -- |
| Monthly Income (in INR) | ≤3,907 | 125 | 13 (10.40) | 112 (89.60) | 0.602 | Reference |
|  | 3,908–11,707 | 34 | 7 (20.59) | 27 (79.41) |  | 0.45 (0.16-1.23); 0.143 |
|  | 11,708–19,515 | 47 | 5 (10.64) | 42 (89.36) |  | 0.96 (0.33-2.90); 1.000 |
|  | 19,516–29,199 | 48 | 3 (6.25) | 45 (93.75) |  | 1.74 (0.47-6.40); 0.561 |
|  | 29,200 –39,032 | 42 | 6 (14.29) | 36 (85.71) |  | 0.70 (0.25-1.97); 0.575 |
|  | 39,033–78,062 | 81 | 8 (9.88) | 73 (90.12) |  | 1.06 (0.42-2.68); 0.903 |
|  | ≥78,063 | 59 | 8 (13.56) | 51 (86.44) |  | 0.74 (0.29-1.90); 0.529 |
|  | Prefer not to say | 26 | 4 (15.38) | 22 (84.62) |  | -- |
| Socioeconomic scale | Upper (I) | 81 | 9 (11.11) | 72 (88.88) | 0.047 | Reference |
|  | Upper Middle (II) | 236 | 22 (9.32) | 214 (90.67) |  | 0.82 (0.36-1.87); 0.640 |
|  | Lower Middle (III) | 36 | 10 (27.77) | 26 (72.22) |  | 3.07 (1.13-8.41); 0.024 |
|  | Upper Lower (IV) | 108 | 13 (12.03) | 95 (87.96) |  | 1.09 (0.44-2.70); 0.844 |
|  | Lower (V) | 1 | 0 | 1 (100) |  | -- |

## Discussion

This study presented the information on knowledge and beliefs of general public of India on COVID-19. To the best of our knowledge this is the first Indian on this hypothesis. Participants from all adult age categories, education, occupation and socioeconomic status participated in this survey. Despite many educational programmes initiated by the government and private organizations on prevention of COVID-19, it is noteworthy that the considerable fraction of participants were unaware of all risk factors and modes of transmission of new coronavirus. This fraction of participants could be potential vectors for spread of infection especially in a pandemic like COVID-19. Educating people on survival time of new coronavirus on various objects would help people to understand the need of disinfecting the objects. Creating awareness among people on individuals who are at risk of acquiring COVID-19 will help people to keep themselves in self-quarantine and simultaneously helps in further spread of infection. Participants were aware of major symptoms and steps to be taken if they were in contact with COVID-19 patient. From this study it was identified that almost equal number of participants relied on news channels followed by healthcare professionals, government websites and social media such as WhatsApp, Facebook for the information on COVID-19. However, the credibility of information being received from social media is still debatable.

Regarding beliefs of participants on COVID-19, mixed type of responses were observed in this section. Although majority of participants were aware of myths and facts on COVID-19, noticeable number of participants were believing some of these myths. More than 50% of participants were unaware or not sure if UV disinfection lamps, swallowing or gargling warm water and eating garlic helps in prevention or treating COVID-19. According to WHO UV lamps, swallowing or gargling warm or saltwater and consuming garlic will not kill the new coronavirus.^[15]^ Furthermore, WHO stated that use of UV lamps for sterilizing hands may cause skin irritation.

Pandemics does not follow the usual pattern as we see in normal infections. There are no studies conducted stating that changes in temperature or taking hot water bath will kill the new coronavirus. However, based on the study conducted on one of the strains of coronavirus i.e SARS, the maximum survival temperature is 56 °C, which is higher than the normal body temperature (36.5 to 37 °C).^[19]^ The new coronavirus affects the respiratory system of the healthy individual who inhale the virus contained droplets or contacted the contaminated surfaces. According to WHO there is no information or studies till date stating that the new coronavirus can be transmitted through mosquitos.^[15]^ Alcohol and bleach are strong and effective disinfectants. Alcohol is a highly flammable compound, it is been used to disinfect small surfaces in ventilated places. However, there is no evidence stating that drinking alcohol will kill the new coronavirus. Furthermore, excessive consumption of alcohol may result in serious health complications.^[20]^ Bleach is easily available at low costs. It is effective against bacteria, fungi and viruses including influenza virus. But it easily reacts with other chemicals and decomposes in presence of heat and light. Contacting with bleach or spraying it over the body would irritate the skin and mucous membrane.^[20]^

This study identified that the people who are aged, having low level of education and non-professionals are having more of false belief towards transmission of new coronavirus, and prevention & treatment of COVID-19. Therefore, it is suggested that the governments or non-government organizations should have a special focus on educating these categories of population. Furthermore, it is not only the duty of government on educating the people, it is the duty of every citizen of India to educate themselves and others on facts of COVID-19 by relying on trusted sources.

## Conclusion

This study was assessed the knowledge and beliefs of general public of India on COVID-19, and also the factors which are influencing the beliefs of participants. As per the observations in this study, the effort of government agencies of providing accurate information on preventive strategies of COVID 19 has shown a greater impact on the knowledge components about COVID-19. However, still there is need for combined efforts of governments and non-government organizations to educate the people to avoid the false beliefs especially among the people who are elderly, having low level of education and non-professional workers on various aspects of COVID-19.

## Data Availability

Complete data of this manuscript is available with the authors.

## Acknowledgement

Authors would like to thank Dr. M Ramesh, Professor & Head, Dept. Pharmacy Practice, JSS College of Pharmacy, JSS AHER, Mysuru and Dr. T M Pramod Kumar, Principal, JSS College of Pharmacy, JSS AHER, Mysuru for permitting us to conduct this study, and also we thank all respondents for sparing their time in filling the questionnaire.

